# A preliminary study on analytical performance of serological assay for SARS-CoV-2 IgM/IgG and application in clinical practice

**DOI:** 10.1101/2020.05.05.20092551

**Authors:** Quan Zhou, Danping Zhu, Huacheng Yan, Jingwen Quan, Zhenzhan Kuang, Weiyun Zhang, Ling Huang, Ting Lei, Jiahui Liu, Bin Xiao, Aiwu Luo, Zhaohui Sun, Linhai Li

## Abstract

**Objective:** To investigate the performance of serological test and dynamics of serum antibody with the progress of SARS-CoV-2 infections.

**Methods:** A total of 419 patients were enrolled including 19 confirmed cases and 400 patients from fever clinics. Their serial serum samples collected during the hospitalization were menstruated for IgM and IgG against SARS-CoV-2 using gold immunochromatographic assay and chemiluminescence immunoassay. We investigated whether thermal inactivation could affect the results of antibody detection. The dynamics of antibodies with the disease progress and false positive factors for antibody testing were also analyzed.

**Results:** The positive rate of IgG detection was 91.67% and 83.33% using two CLIA, respectively. However, the IgM positive rate was dramatically declined might due to the lack of blood samples at early stages of the disease. The chemiluminescence immunoassay had a favorable but narrow linear range. Our work showed increased IgG values in serums from virus-negative patients and four negative samples were IgG weak-positive after thermal incubation. Our data showed the specificity of viral N+S proteins was higher than single antigen. Unlike generally thought that IgM appeared earlier than IgG, there is no certain chronological order of IgM and IgG seroconversion in COVID-19 patients. It was difficult to detect antibodies in asymptomatic patients suggesting that their low viral loads were not enough to cause immune response. Analysis of common interferent in three IgG false-positive patients, such as rheumatoid factor, proved that false positives were not caused by these interfering substances and antigenic cross-reaction.

**Conclusions:** Viral serological test is an effective means for SARS-CoV-2 infect detection using both chemiluminescence immunoassay and gold immunochromatographic assay. Chemiluminescence immunoassay against multi-antigens has obvious advantages but still need improve in reducing false positives.

## Introduction

Coronavirus disease 2019 (COVID-19, formerly known as 2019-nCoV), a new form of respiratory and systemic disorder sustained by the severe acute respiratory syndrome coronavirus 2 (SARS-CoV-2)^1^, which leads to a global serious public health issue especially in Wuhan city, Hubei province, China since December 2019^2^. The majority of COVID-19 patients have an incubation period of 3 to 7 days^3^. Typical symptoms include fever, cough and fatigue^4^, while severe cases might rapidly progress to acute respiratory distress syndrome (ARDS), septic shock and difficult-to-tackle metabolic acidosis and bleeding and coagulation dysfunction^5^. It is evidenced that SARS-CoV-2 can transmit rapidly from person to person through a variety of methods including droplets, contact and aerosols, which is evidently found in hospital and family settings^6-8^. By May 5^th^, there are over 357 million people worldwide have been infected with SARS-CoV-2, with approximately 250,000 deaths.

As a member of betacoronavirus family, SARS-CoV-2 are characterized by a single-stranded positive-sense RNA genome with a size of 29.8 kb^1^. A typical CoV mainly contains four structural proteins: spike (S), membrane (M), envelope (E), and nucleocapsid (N) proteins. The S protein is required for attachment to host receptors^9^. Glycoprotein M is involved in the formation and budding of viral envelopes^10^. Nucleocapsid protein N is the most abundant and conservative structural protein in coronaviruses. After self-polymerization, it coats the viral RNA genome to form a spiral nucleocapsid^11^.

Many domestic institutions, including the Chinese Center for Disease Control and Prevention (CCDC), firstly reported in the New England Journal of Medicine about the comprehensive information of SARS-CoV-2,including virus isolation, structure, genome sequence, and evolution^8^. On the basis of analysis of three complete genomes obtained in this study, the researchers designed several specific and sensitive assays targeting ORF1ab, N, and E regions of the 2019-nCoV genome to detect viral RNA in clinical specimens^12^. Nucleic acid detection is performed by searching for fragments of viral genetic material from nasal and pharyngeal swabs or fluid collected from the lungs, which positive results can be diagnosed as COVID-19 but negative cannot rule out the infection. IgM/IgG antibody detection can quickly detect patient infection and monitor the progress of rehabilitation. SARS-CoV-2 antibody assay kits currently on the market were mainly established for detecting IgM or IgG antibody against N and S1 protein antigens. Despite the shortcomings of “window period” and “crossover antigen”, serological diagnostic methods have become complementary means of nucleic acid testing for SARS-CoV-2 detection.

In the present study, we applied different antibody detection methods to monitor the antibody response to SARS-CoV-2 in 419 patients in fever clinics including 19 COVID-19 patients. We further investigated whether thermal inactivation affected the efficiency of SARS-CoV-2 antibody detection and false positive factors for antibody testing using chemiluminescence immunoassay.

## Methods

### 1. Patients and samples

This study enrolled a total of 419 cases including 19 COVID-19 cases, where all patients were admitted to General Hospital of Southern Theater Command of PLA from January 22 to April 04, 2020. COVID-19 or NON-COVID-19 cases were confirmed to be infected with or without SARS-Cov-2 by real-time PCR. Among the all 419 enrolled patients, 19 were confirmed SARS-Cov-2 infection, who were tested positive for viral RNA using real time RT-PCR assay on pharyngeal swab specimens; the others were reported negative. The general information (age, sex, vital signs, coexisting disorders), clinical and laboratory characteristics data of the patients were collected. The serum was collected from each enrolled patient for antibody test. The study was reviewed approved by the Medical Ethical Committee of General Hospital of Southern Theater Command of PLA.

### 2. Sample dilution and inactivation

The serum samples were serially diluted using saline or virus-negative serum. Two series of dilution were carried out, namely original to 32 times using two different diluent agents. Serum samples were inactivated by incubation in a water bath at 56°C for 60 minutes.

### 3. Instruments and reagents

Virus preservation tube was purchased from Shenzhen Biocomma Biological Science and Technology Co., Ltd. Automatic nucleic acid extraction instrument MFL purifier96 Magnetic particle separator was purchase from Genfine Biological Science and Technology (Changzhou) Co., Ltd. Real time RT-PCR was performed using the nucleic acid testing kit (Daan, Guangzhou, China) for SARS-CoV-2 detection on a Roche LightCycler 480 real-time PCR instrument. The IgM antibody and IgG antibody against SARS-CoV-2 kits (chemiluminescence immunoassay, CLIA) and their testing equipment were purchased from Shenzhen YHLO Biotech Co., Ltd. and Shenzhen Tisenc Medical Devices Co., Ltd.

### 4. Real time Reverse Transcription Polymerase Chain Reaction (RT-PCR) Assay

The SARS-CoV-2 laboratory test assays were based on the guideline of laboratory detection for COVID-19. Pharyngeal swab specimens were obtained from patients and restored into a tube with 200 ul of virus preservation solution. RNA was extracted and tested by real-time RT-PCR with 2019-nCoV–specific primers and probes followed manufacturer’s instructions without modification. Briefly, RNA was extracted from 200 ul Virus preservation solution, Real-time PCR was conducted using the nucleic acid testing kit (Huada, Guangzhou, China) in a Roach LC480 System with the manufacturer’s protocol.

### 5. Chemiluminescence immunoassay and colloidal gold immunochromatography test

To quantitatively detect anti-SARS-Cov-2 antibodies in serum, two chemiluminescence immunoassays (CLIA) were applied. The commercially-available single-assay chemiluminescence test (Yahuilong) which was developed using SARS-Cov-2 S protein and N protein as antigens was conducted in a Yahuilong iFlash 3000-H System according to the manufacturer’s instructions. Cutoff value was calculated from relative luminescence units (RLU) and calibrator standard curve, the cutoff value was 10 AU/mL. RLU less than cutoff value was defined as negative, and RLU greater than or equal to cutoff value was defined as positive. Another test reagent was purchased from Shenzhen Tisenc Medical Devices Company. All operations were performed in the ACCRE 6 system according to the instructions.

The antibody qualitative test was also carried out using gold immunochromatographic assay (GICA) purchased from Guangzhou Wondfo company. All serum samples were collected to detect antibody against SARS-Cov-2 virus by gold immuo-chromatographic assay according to the manufacturer’s operation manual.

### 6. Statistical analysis

GraphPad Prism Version 8 software was employed for statistical analysis. The categorical variables were expressed as the counts and percentages and compared using Fisher’s exact test.

Continuous variables were expressed as the mean±SD or median with interquartile ranges (IQR). Mann-Whitney U test was used to compare the quantitative serological assay results. To check out the effect of inactivation on antibody concentration measurement, paired t-test was carried out. Spearman correlation analysis was employed to analyze the correlation between groups. Differences were considered significant at p< 0.05 with a two-tail test.

## Results

### 1. Demographic characteristics of cases

As shown in table 1, 19 COVID-19 patients and 400 Non-COVID-19 patients who were treated in fever clinics and wards of General Hospital of Southern Theater Command of PLA from January 22 to April 04, 2020, were enrolled in this study.

**Table 1.**
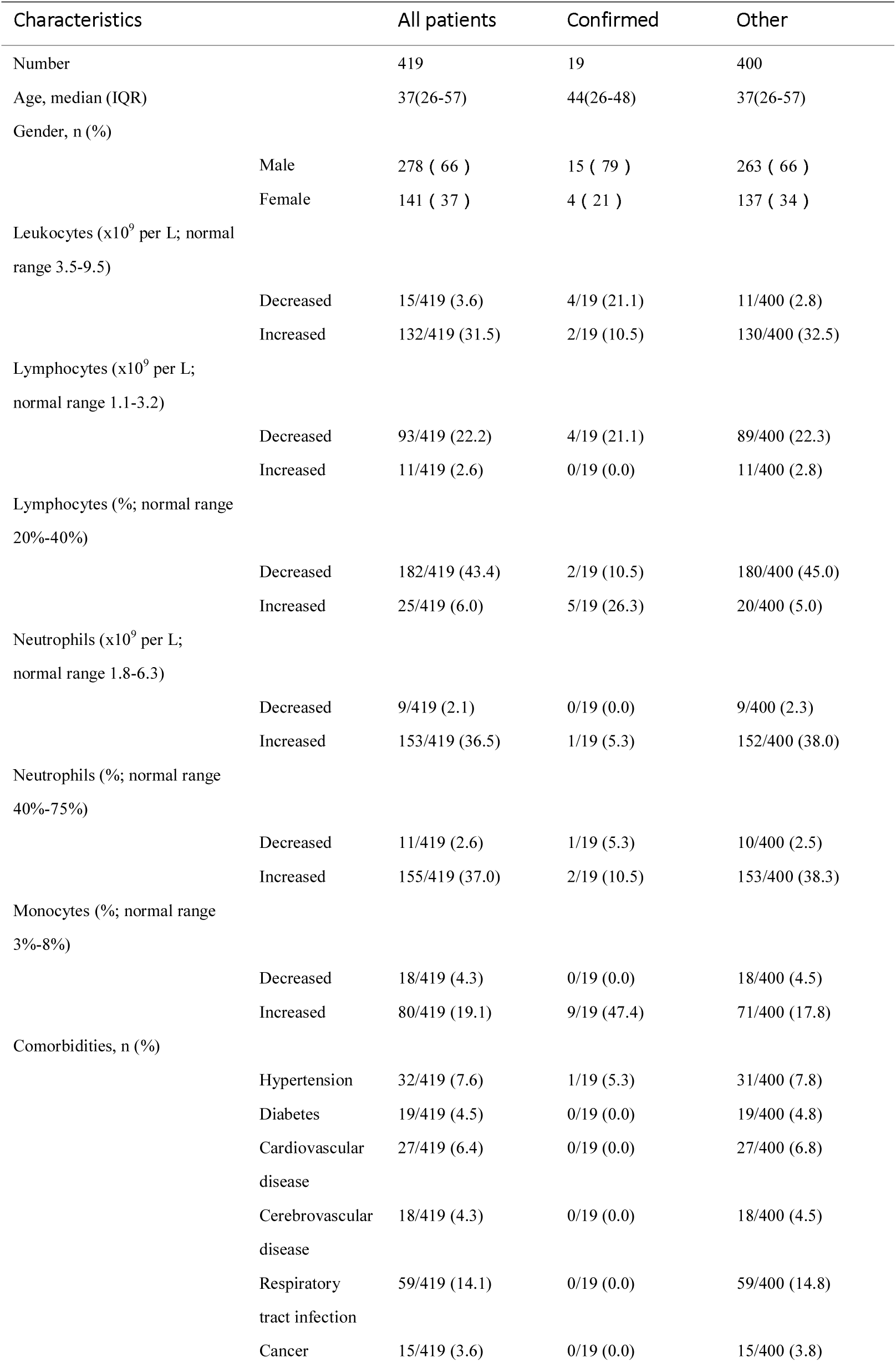
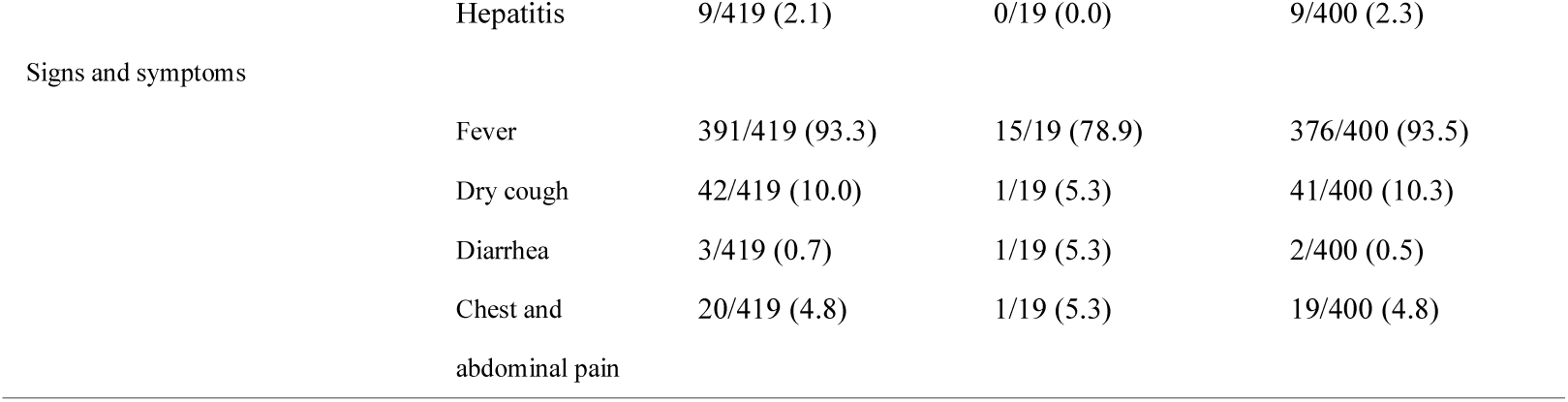
Characteristics of 419 enrolled patients

The median age was 37 years (IQR, 26-57) and the proportion of men was 66%. Among these patients, the most common symptoms were fever (391, 93.3%), dry cough (42, 10%) and Chest and abdominal pain (20, 4.8%). Respiratory tract infection (59, 14.1%), hypertension (32, 7.6%), cardiovascular disease (27, 6.4%) and diabetes (19, 4.5%) were the most common coexisting comorbidities. Other comorbidities on admission were cerebrovascular disease (18, 4.3%), cancer (15, 3.6%) and hepatitis (9, 2.1%).

At illness onset, 93 (22.2%) patients had lymphocytes below the normal range and 11 (2.6%) patients were found above the normal range. Leucocytes were below the normal range in 15 (3.6%) cases and above the normal range in 132 (31.5%) cases. There were 9 (2.1) patients had neutrophils below the normal range and 153 (37.0%) above. The percentage of monocytes was increased in 80 (19.1%) patients and decreased in 18 (4.3) patients. A total of 946 blood sample and 463 pharyngeal swab specimens were collected.

### 2. Performance of CLIA for viral antibody detection

To validate the performance of CLIA (Yahuilong), a series of dilution experiments were performed using serum samples from 3 confirmed COVID-19 patients with saline or virus-negative serum. We first measured the IgG CLIA dilution assay with virus-negative serum in a narrow limit which serially diluted by a factor of 2 to 32 times. The line chart presented the dilution curve of viral IgG concentration of each specimen (Fig. 1 A). As the figure shown, the CLIA had a good linear range which correlation coefficient was greater than 0.9 for IgG detection. Then the dilution assay was carried out using saline. The correlation coefficients were also greater than 0.9 but increased compared using serum ones (Fig. 1 B). These results suggested that both saline and virus-negative serum can be used as diluent reagent in CLIA but saline may have minor interference. We also observed that when the value reached about 70 AU/mL the increment no longer increased proportionally. If only using the data points less than 70 AU/mL to calculate the linear coefficient, it dramatically increased to 0.99 suggesting a narrow linear range.

**Figure 1.**
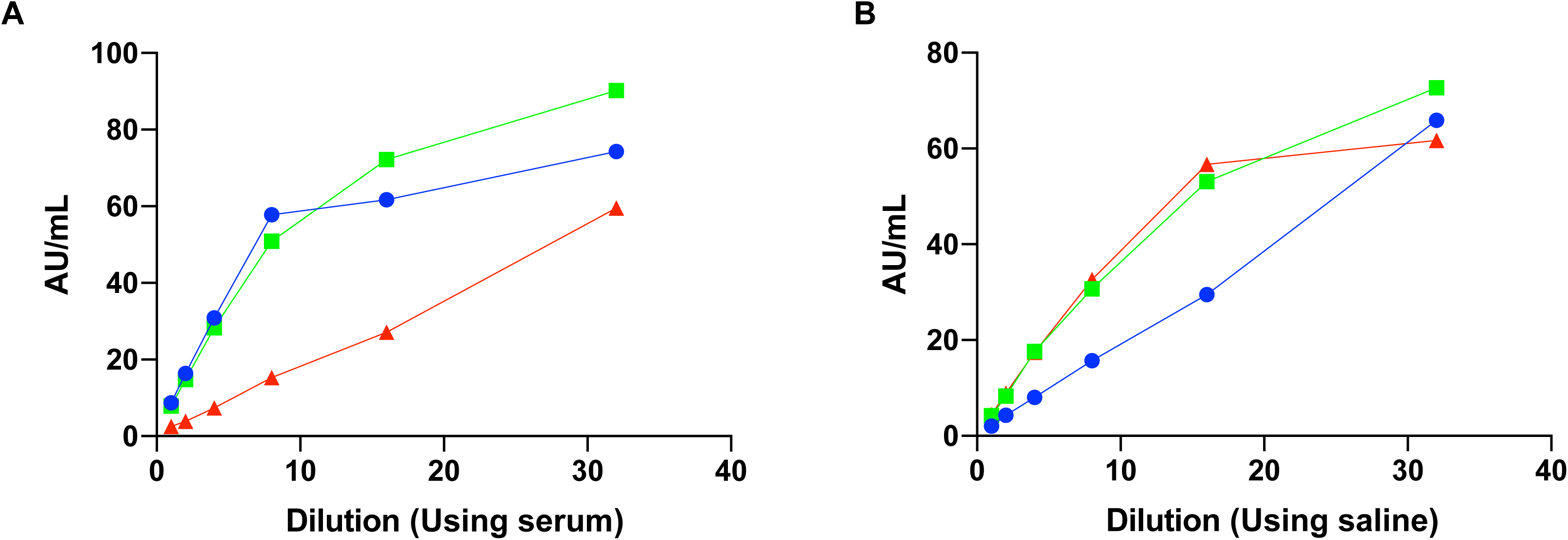
IgG antibody assay linearity results using negative-serum (A) and saline (B) diluent. Six virus-positive serum samples were serially diluted by a factor of 2 to 32 times. (A) The correlation coefficient of three curves is 0.86, 0.94 and 0.99, respectively. (B) The correlation coefficient of three curves is 0.99, 0.97 and 0.92, respectively.

To determine the impact of thermal inactivation, parallel CLIA was performed using serum from 397 ordinary patients and 10 confirmed COVID-19 patients. The line chart presented contrasts the antibody concentration with or without incubation at 56°C for 60 min (Fig. 2). For the virus-negative serum, there were no difference in concentration mensuration of IgM between the inactivated and original samples and didn’t affect the qualitative results (Fig2.A). However, there was a significant difference in IgG mensuration, the antibody concentrations menstruated in inactivation group were higher than the original groups (Fig.2 B). Several virus-negative patients had seropositivity IgG antibodies, which present they were false-positive or individuals previously infected. Notably, negative results in 4 cases which below the critical value were converted into false positives after inactivation, which indicated the inactivation operation may impair the decision outcome especially the concentration was near the cutoff value (Fig2 B). In the confirmed cases group, the mensuration of both IgM and IgG had no significant difference with or without inactivation. Moreover, unlike the negative group, the inactivation didn’t have an impact on qualitative results (Fig.2 C and D).

**Figure 2.**
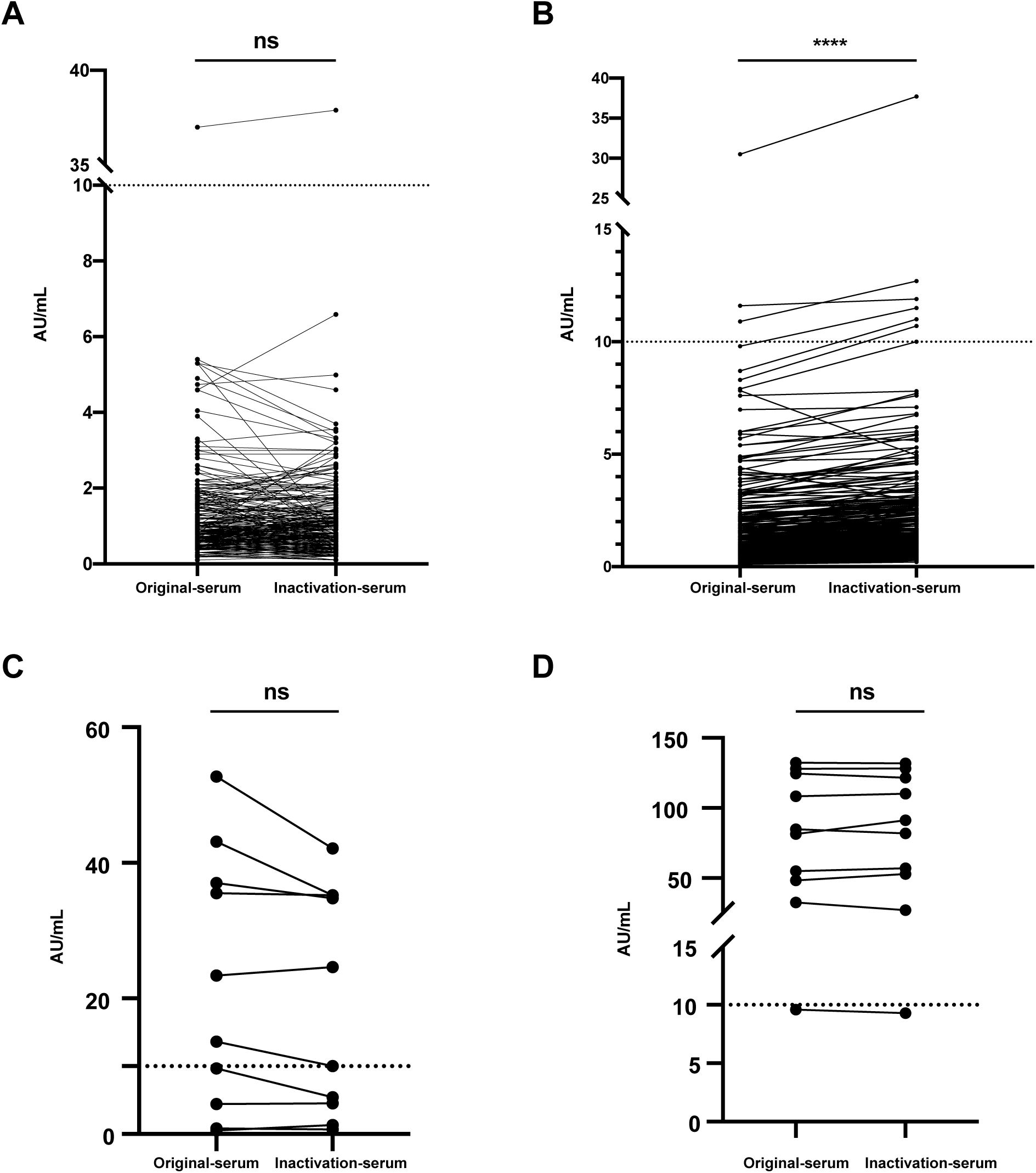
Effects of thermal inactivation on the IgM/IgG antibody tests of SARS-CoV-2 negative (A and B) or positive (C and D) serum samples. Each sample was detected with or without incubation at 56°C for 60 minutes. Each dot represents the measured value by CLIA and dotted line represents the cutoff value namely 10 AU/mL. Virus-negative samples (n=397) were tested for IgM (A) and IgG (B). Virus-positive samples (n=10) were tested for IgM (C) and IgG (D). The comparison between original and inactivation groups was tested by paired two-tailed Student’s f-test. ^***^ represents *P*<0.001.

We also compared different methods from various manufacturers to detect antibodies in ten COVID-19 patients, including quantitative CLIA purchased from Yahuilong and TISENC companies, qualitative GICA purchased from Wondfo company. As shown in Table 2, the positive rate of IgG detection was 91.67% and 83.33% using two CLIA, respectively. The false-negative patients were asymptomatic. However, the IgM positive rate was dramatically declined in both two approaches. This may be caused by the middle and late stages of the disease. The IgG false positive rate of two methods was 1.25% and 0.76% respectively.

**Table 2.** Performance of different detection methods

|  | Viral RNA | IgG ( Yahui long ) | IgM ( Yahui long ) | IgG<br>( TISENC ) | IgM<br>( TISENC ) | Ab ( Wondfo ) |
| --- | --- | --- | --- | --- | --- | --- |
| Methods | qPCR | Indirect CLIA | Indirect CLIA | Indirect CLIA | Indirect CLIA | Capture GICA |
| Targets | N+ORF1a/b | N+S1 | N+S1 | S | S | S1 |
| Matched cases | 12/12 | 11/12<br>( 91.67% ) | 4/12<br>( 33.33% ) | 10/12<br>( 83.33% ) | 1/12<br>( 8.33% ) | 10/12<br>( 83.33% ) |
| Negative | 0/400 | 5/400<br>( 1.25% ) | 1/400<br>( 0.25% ) | 3/392<br>( 0.76% ) | 0/392<br>( 0.0% ) | 0/400<br>( 0.0% ) |

Confirmed cases were based on qPCR viral RNA detection.

For qualitative method, GICA was to detect total antibodies against virus S protein. The dipstick had a relatively lower positive rate (83.3) and no false-positive case reported. Notably, the two patients (case 17 and 18) had negative Ab colloidal gold test strip results were asymptomatic infected. Their quantitative results detected by CLIA were negative, either.

These results indicated that these two methods were feasible in qualitative and quantitative detecting antibodies. Subsequent experiments were carried out using chemiluminescence on the iFlash CLIA platform to determine antibody concentration.

### 3. The dynamics of antibody levels with the progress of virus infections

To investigate the dynamics of antibody levels with disease progression, we conducted long-term antibody level monitoring on five representative confirmed cases including one asymptomatic patient. The first positive time point of Ab tests appeared later than that of RNA in 4 of 5 patients as expected, except for case 8. Ab was detectable at the same day with RNA detected of case 8, and both IgM and IgG were maintained at a high level in the first two weeks. The patient reported that symptoms such as fatigue and chills had occurred one week before the diagnosis, suggesting that it had been infected for a period of time. More surprisingly, the patient’s antibody levels were remained above-normal values in almost three months since illness onset (Fig. 3A).

**Figure 3.**
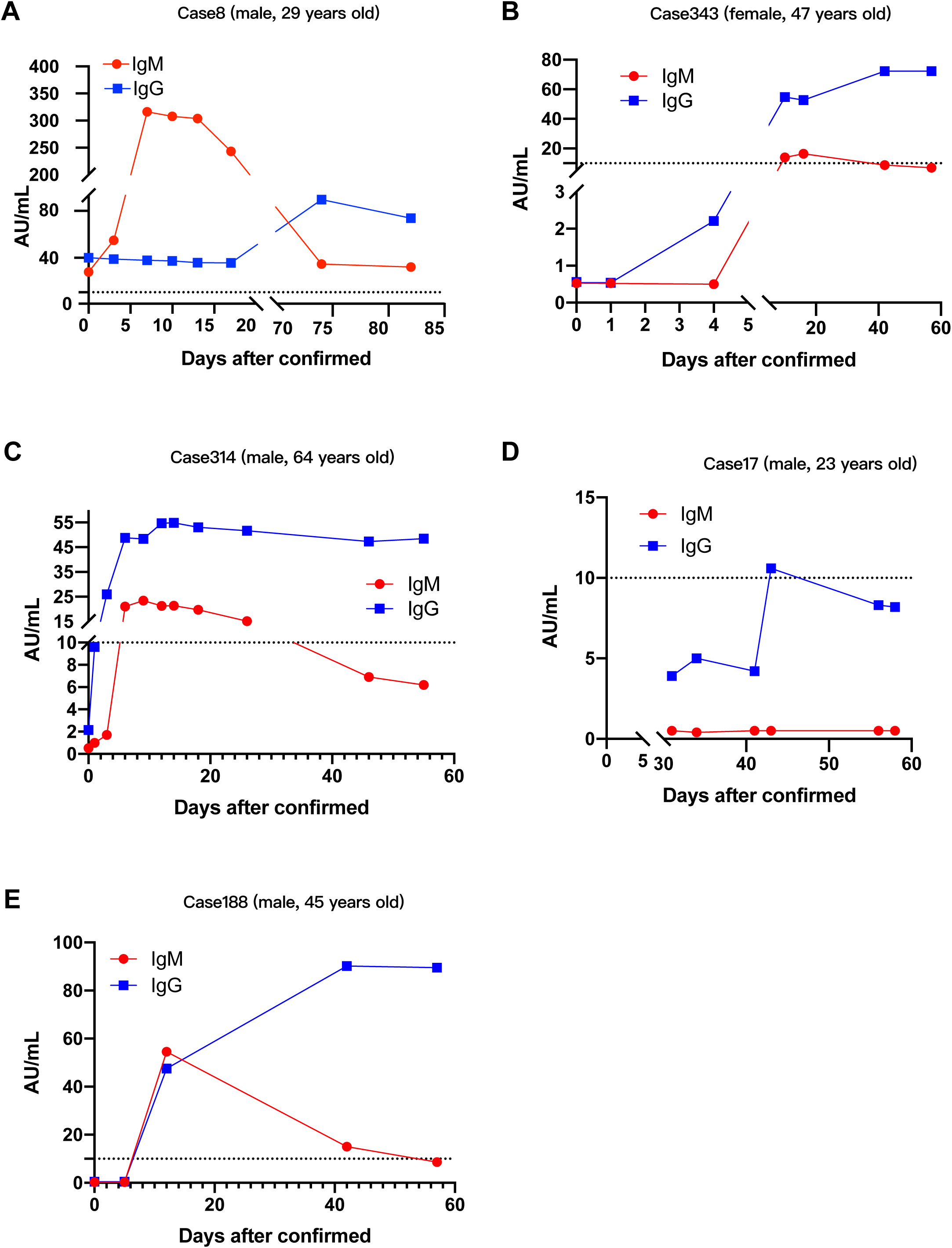
Temporal dynamic profiles of serum IgM and IgG against viral N+S proteins, as ascertained by CLIA. Among the 19 patients, 5 achieved seroconversions in the observation. The red dots and blue squares represent the seroconversion points of IgM and IgG respectively. Dotted line represents the cutoff value.

The level of Ab was constantly below the cutoff value in case 17 except only one IgG point and rapidly decreased later (Fig. 3D). But the patient started antibody monitoring nearly one month after diagnosis, the IgM may have declined. It is worth noting that case 17 was also an asymptomatic patient. It seems these asymptomatic patients had low viral loads not enough to cause immune response. But these conclusions still need to be further verified in a large asymptomatic patient cohort.

It is generally believed that IgM appears before IgG in the early stage of infection. But among the 5 patients we monitored, the first positive time point of IgG tests appeared earlier than IgM in 1 patient (case 314), and 1 patient (case 188) reported positive results of IgG and IgM simultaneously (Fig 3 C and E).

These results indicated diagnosing the course of COVID-19 based on the time when the antibody appeared was unreliable in some patients and the dynamics of antibody response in COVID-19 may be different from previous infections. However, due to insufficient sample size, whether these conclusions were general or exceptional needs further verification.

### 4. False positive factors for CLIA antibody testing

There were 5 seropositive patients of 400 nucleic acid-negative patients, three of them were reported antibody positive with both two CLIA assays but negative in immunochromatographic test strip (Table 2). We analyzed these three samples to explore false positive factors. First, we confirmed the positive value was indeed an immune binding signal through dilution experiments, not other unrelated interference signals (Fig.4 A). Rheumatoid factor (RF) is a common interference factor for the determination of antibodies by chemiluminescence, so we conducted RF measurements on these three samples. The results showed that the total RF antibodies, IGG and IGM were all negative, so RF was not the main interference factor of false positive (Table 3). Then we checked whether the false positives were caused by reagents. Different batches and manufacturer R&D reagents were used for test. To some extent, the false positive signal value decreased, but it did not turn negative in two cases (Fig.4 B). Next, we tested whether adding blockers could eliminate false positives. We added heterophilic antibody blockers and commercial blockers to serum diluents then detected antibody RLU. The three diluents adding heterophilic antibody blockers showed slightly lower RLU than those without blockers, but there were still 2 samples with positive results; adding commercial blockers had no effect on the measuring results (Fig.4 C). These data mentioned above indicated there were positive signals related to the homology of antigen fragments in false positive serums, and it is necessary to optimize the selection of antigen fragments to improve the specificity.

**Table 3.** RF testing results of three false-positive cases

| Case | RF<br>( cutoff = 3 ) | RF IgG<br>( cutoff = 4 ) | RF IgM<br>( cutoff = 10 ) |
| --- | --- | --- | --- |
| Case 308 | Negative | Negative | Negative |
| Case 385 | Negative | Negative | Negative |
| Case 363 | Negative | Negative | Negative |

**Figure 4.**
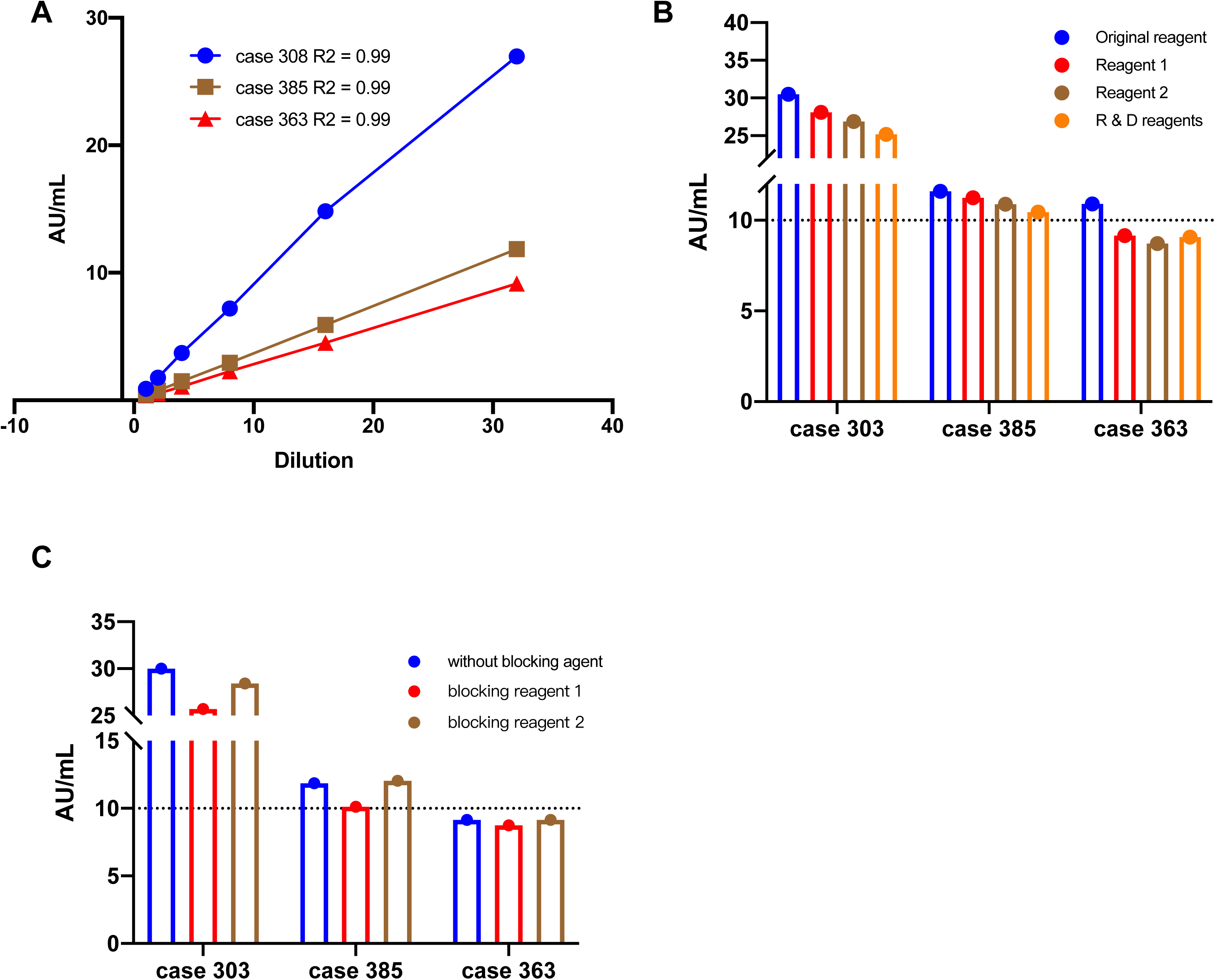
Exploring false positive factors for CLIA antibody testing. (A) Dilution curves for three IgG false positive cases. All curves showed favorable correlation coefficients indicating the signals were indeed caused by immune binding. (B) Reagents of different batches were used for IgG detection to exclude the batch effect. (C) Measuring IgG antibody after adding heterophilic antibody blockers (reagent 1) and commercial blockers (reagent 2). Dotted line represents the cutoff value.Tables

## Discussion

The rapid spread of COVID-19 represents a major challenge for public health systems worldwide. In addition to treating patients and preventing the spread of disease, laboratory diagnosis is also a critical part for curbing outbreaks. Viral nucleic acid detection was the gold standard for COVID-19 confirmation as recommended, but with the improvement of various antibody detection methods, the antibody detection has risen in importance unceasingly. According to Chinese Clinical Guidance for COVID-19 Pneumonia Diagnosis and Treatment (7^th^ edition), serological testing, including gold immunochromatographic assay (GICA), enzyme-linked immunosorbent assay (ELISA) and chemiluminescence immunoassay (CLIA), were the diagnostic criteria for confirmed cases and the exclusion criteria for suspected cases. Among them, GICA is simple and quick, and can perform qualitative detection of antibodies; ELISA and CLIA can perform automated quantitative analysis of antibodies. In this study, we preliminarily compared the performance of different methods and manufacturers in detecting SARS-Cov-2 antibodies. The positive rate of most reagents was about 90% or above, but the performance of detecting IgM was unsatisfactory, probably because its concentration was too low to detect, and that IgM rose then decreased rapidly in early and mid-infection. Both CLIA used indirect methods to measure antibodies, but the coated antigens were different (Table 2). Our data showed the specificity of viral N+S proteins was higher than single antigen. So, we recommend the assay had coated multi-antigens. However, neither method can detect antibodies in two cases of asymptomatic infections, suggesting that for asymptomatic infections, the lower viral load cannot trigger sufficient immune response.

Owing to the contagiousness of SARS-CoV-2, thermal inactivation has been recommended before laboratory testing to reduce the risk of viral transmission. A previous study indicated thermal inactivation could result in a decreased detectable amount of viral nucleic acid and increased Ct values in RT-PCR detection which caused false-negative results^13^. In our study, it was found that in some serum of patients with negative nucleic acids test, the antibody concentration increased after inactivation, which will cause false positive. When performing antibody test with thermal inactivation, it should be very careful with the sample around cut-off value to avoid false positive results.

The time kinetics of IgM and IgG was also evaluated. Generally, both IgM and IgG rapidly increased after the onset of fever. However, as mentioned above, the antibody levels of two asymptomatic patients were below the cutoff value in the course of disease.

Zhang et al. found that the increase of antibody was clearly visible in almost all patients after 5 days of symptom onset, which considered as a period of time from early to mid-term infection^14^. Unlike generally thought that IgM appeared earlier than IgG, there is no general rule for the chronological order of IgM and IgG seroconversion for a specific patient, which resembles the conditions in SARS^15^ and MERS^16^ and also observed in SARS-CoV-2 by other researchers^17^. Therefore, we recommend using total antibodies instead of measuring IgM and IgG alone to determine the progress of infection and diagnosis.

False positive is one of the common problems in chemiluminescence immunoassay. In our study, there were five IgG positive cases among 400 nucleic acid negative patients which were considered false positive. Analysis of common interferent, such as rheumatoid factor, proved that false positives were not caused by these interfering substances. The results suggested that further optimization was needed to improve the detection specificity.

The present study has some notable limitations. First, only 19 laboratory confirmed COVID-19 patients were included. It can’t tell whether certain conclusions were exceptional or universal. Second, as a retrospective study, it was difficult to completely match the sampling time and detection time. Therefore, in antibody dynamic monitoring, there was large time spans resulting in incomplete dynamic curves. Third, viral serological detection kits were available late after COVID-19 outbreak, and IgM detection in the acute phase will be partially lost. Finally, due to the limitation of the sample size and serum amount, no systematic analysis has been carried out in comparison of various reagents, and the conclusion may be biased.

In conclusion, viral serological testing is an effective means for SARS-CoV-2 infection detection using both chemiluminescence immunoassay and gold immunochromatographic assay.

## Data Availability

All data used during the study are available from the corresponding author by request.

## Reference

1. Lu R, Zhao X, Li J, et al. Genomic characterisation and epidemiology of 2019 novel coronavirus: implications for virus origins and receptor binding. The Lancet 2020; 395(10224): 565–74.

2. Huang C, Wang Y, Li X, et al. Clinical features of patients infected with 2019 novel coronavirus in Wuhan, China. The Lancet 2020; 395(10223): 497–506.

3. Guan W, Ni Z, Hu Y, et al. Clinical characteristics of 2019 novel coronavirus infection in China. *medRxiv* 2020.

4. Wang M, Wu Q, Xu W, et al. Clinical diagnosis of 8274 samples with 2019-novel coronavirus in Wuhan. *medRxiv* 2020.

5. Chen N, Zhou M, Dong X, et al. Epidemiological and clinical characteristics of 99 cases of 2019 novel coronavirus pneumonia in Wuhan, China: a descriptive study. The Lancet 2020; 395(10223): 507–13.

6. Chan JFW, Yuan S, Kok K, et al. A familial cluster of pneumonia associated with the 2019 novel coronavirus indicating person-to-person transmission: a study of a family cluster. The Lancet 2020; 395(10223): 514–23.

7. Wu JT, Leung K, Leung GM. Nowcasting and forecasting the potential domestic and international spread of the 2019-nCoV outbreak originating in Wuhan, China: a modelling study. The Lancet 2020; 395(10225): 689–97.

8. Li Q, Guan X, Wu P, et al. Early Transmission Dynamics in Wuhan, China, of Novel Coronavirus–Infected Pneumonia. The New England journal of medicine 2020.

9. Delmas B, Laude H. Assembly of coronavirus spike protein into trimers and its role in epitope expression. Journal of Virology 1990; 64(11): 5367–75.

10. Neuman BW, Kiss G, Kunding AH, et al. A structural analysis of M protein in coronavirus assembly and morphology. Journal of Structural Biology 2011; 174(1): 11–22.

11. Hurst KR, Koetzner CA, Masters PS. Identification of In Vivo-Interacting Domains of the Murine Coronavirus Nucleocapsid Protein. Journal of Virology 2009; 83(14): 7221–34.

12. Zhu N, Zhang D, Wang W, et al. A Novel Coronavirus from Patients with Pneumonia in China, 2019. The New England journal of medicine 2020; 382(8): 727–33.

13. Pan Y, Long L, Zhang D, et al. Potential false-negative nucleic acid testing results for Severe Acute Respiratory Syndrome Coronavirus 2 from thermal inactivation of samples with low viral loads. Clinical Chemistry 2020.

14. Zhang W, Du R, Li B, et al. Molecular and serological investigation of 2019-nCoV infected patients: implication of multiple shedding routes. Emerging microbes & infections 2020; 9(1): 386–9.

15. Hsueh P, Huang L, Chen P, Kao C, Yang P. Chronological evolution of IgM, IgA, IgG and neutralisation antibodies after infection with SARS-associated coronavirus. Clinical Microbiology and Infection 2004; 10(12): 1062–6.

16. Drosten C, Meyer B, Muller MA, et al. Transmission of MERS-coronavirus in household contacts. The New England journal of medicine 2014; 371(9): 828–35.

17. Long Q, Deng H, Chen J, et al. Antibody responses to SARS-CoV-2 in COVID-19 patients: the perspective application of serological tests in clinical practice. *medRxiv* 2020.

